# Global incidence of sudden cardiac arrest in young athletes and military members: a systematic review and meta-analysis

**DOI:** 10.1101/2020.09.13.20193714

**Authors:** Aaron Lear, Niraj Patel, Chanda Mullen, Marian Simonson, Vince Leone, Constantinos Koshiaris, David Nunan

**Affiliations:** Cleveland Clinic Akron General, Akron, OH USA; HeyDoctor by GoodRx, San Francisco, CA USA; Cleveland Clinic Foundation, Cleveland, OH USA; Northeast Ohio Medical University (NEOMED), Akron, OH USA; University of Oxford, Oxford, UK

**Author notes:** Contact author: Aaron Lear, 1 Akron General Way, Ctr for Fam Medicine, Akron, OH 44307 USA, (Angela Watkins assistant).

**Keywords:** athletes, military, sudden cardiac arrest, sudden cardiac death, incidence

## Abstract

**Objectives:** The goals of this review are to evaluate the quality of the evidence on the incidence of sudden cardiac arrest and death (SCA/D) in athletes and military members; and to estimate annual incidence of SCA/D.

**Data Sources:** MEDLINE, Embase, Cochrane CENTRAL, Web of Science, BIOSIS, Scopus, SPORT discus, PEDro, and clinicaltrials.gov were searched from inception to dates between 2/21/19 and 7/29/19.

**Study Selection:** Studies which reported incidence of SCA/D or both in athletes, or military members under age 40 were eligible for inclusion. 40 studies were identified for inclusion

**Data Extraction:** Risk of bias was assessed using a validated, customized tool for prevalence studies in all included studies. 12 were found to be low ROB, with the remaining 28 moderate or high ROB. Data was extracted for narrative review, and meta-analysis.

**Data Synthesis:** Random-effects meta-analysis was performed in studies judged to be low risk of bias in two separate categories: 5 studies on regional or national level data including athletes at all levels, and both sexes included 130 events of SCD, with a total of 11,272,560 athlete years showing a cumulative incidence rate of 0.98 [95%CI: 0.62, 1.53] per 100,000 athlete years, with high heterogeneity with *I*^2^ of 78%; 3 Studies focusing on competitive athletes between the ages of 14 and 25 were combined, and included 183 events, and 17,798758 athlete years showing an incidence rate of 1.91[95%CI: 0.71; 5.14] per 100,000 athlete years with high heterogeneity with *I*^2^ of 97%.

**Conclusion:** The worldwide incidence of SCD is a rare event. Low risk of bias studies indicate levels of SCD to be below 2 per 100,000 athlete years. Overall, the quality of the evidence available on the subject of SCA/D is low, but there are high quality individual studies to inform the question of incidence levels.

**PROSPERO Registration:** CRD42019125560

**Key Points:** There are several published articles which give a clear picture on the estimate of sudden cardiac arrest and death in athletes and military members, but the overall state of the literature has substantial risk of bias, with only 12 of 40 included articles at low risk of bias.

Meta-analysis of sudden cardiac death was shown to be rare overall in athletes, with high quality, large population level studies synthesized to show a rate of 0.98 [95%CI: 0.62, 1.53] per 100,000 athlete years, and more focused studies on competitive younger athletes synthesized to show a rate 1.91[95%CI: 0.71; 5.14] per 100,000 athlete years. There was high heterogeneity present in both meta-analyses.

## Introduction

Sudden cardiac arrest (SCA) leading to death has been shown to be the leading medical cause of death in young competitive athletes^1^. It is believed that exercise and physical activity amongst athletes can lead to episodes of arrhythmia leading to sudden death^2^. The topic of screening athletes with electrocardiograms in order to prevent sudden cardiac arrest and death (SCA/D) has become a controversial topic in sports medicine^3,4^. Authors have pointed out that the overall burden of sudden cardiac death (SCD) remains low, around 1 event per 100,000 athletes/year or less ^3^, while others believe the incidence of SCA/D to be chronically undercounted ^5^, and suggest that screening may decrease the incidence of SCA/D. A lack of consensus on methods to attribute sudden deaths to cardiovascular causes further confuses the issue. Surveys of high schools and universities ^6,7^; search of newspaper and web based reports ^8,9^; use of national health and autopsy records ^10–12^; catastrophic insurance records ^13,14^; and reporting on non-traumatic deaths in the military ^15–17^ have all been performed to determine incidence rates of SCA/D. Resulting estimates have varied widely, and whilst the validity of these different attempts has been questioned^3,4,18^, existing data and their quality have not been adequately assessed systematically.

One systematic review has previously been published on the incidence of ‘sports related’ sudden cardiac death ^19^. The review was not pre-registered, did not report its methods according to existing reporting guidelines and assessment of the quality of evidence was not performed, resulting in more questions than answers. The United Kingdom (UK) National Screening Committee has reviewed data on incidence and screening in the young, and produced results recommendations for practice in the UK^20^. We are not aware of a comprehensive, pre-registered, peer-reviewed systematic review of the evidence base for the incidence of SCA and SCD in young people regularly participating in regular exercise. Understanding the existing evidence and it’s quality on the epidemiology of SCA and SCD is important when considering professional body recommendations^18,21–25^ for screening athletes with electrocardiograms (ECG) for conditions which may cause such events.

Our objective is to identify publications reporting on the incidence of SCA/D in athletes and military members 40 years of age or younger, assess the quality of the evidence, and synthesize a population level incidence of both SCA and SCD.

## Methods

This review is one portion of a systematic review with two objectives: Identifying the incidence of SCA and SCD in young athletes and military members; and the effect on SCA/D of ECG screening this population^26^. The project was performed according to the Preferred Reporting Items for Systematic Reviews and Meta-Analyses (PRISMA)^27^ guidelines, and was registered on PROSPERO March 18^th^ 2019 (CRD42019125560) ^28^.

### Data Sources and Searches

The search strategy was designed with a medical librarian (MS) and combined the dual objectives of the project into a single search. Searches were performed in MEDLINE, Embase, Cochrane CENTRAL, Web of Science, BIOSIS, Scopus, SPORT discus, PEDro, between 2/21/19 and 3/1/19, and Clinicaltrials.gov on 7/29/19 for available articles. Relevant review articles and position statements were hand searched for eligible articles ^5,18,19^, and articles not identified in the online search which reported incidence rates were selected for screening. There was no limitation on language of publication or date. Conference abstracts, published abstracts, and full text publications were included in the review.

### Study Selection

Cohort studies, historically controlled trials, and survey studies which reported incidence of SCA, combining both incidents of SCA and SCD or SCD alone in athletes, or military members age 40 and younger, were eligible for inclusion. Studies which did not report the incidence rate as the primary outcome were included if they reported a rate, or data from which an incidence rate could be calculated. In cases where the study reported events confirmed as SCA or SCD, and suspected cases of SCA or SCD, only the confirmed cases were included in data syntheses. In cases where the authors used a multiplier to address concerns about either cases of SCA/D (numerator), or those eligible for inclusion (denominator), we elected to use the number presented by the authors with the multiplier. This formula was used to calculate either the numerator or denominator when the number of cases or person years was not explicitly detailed: (total number of events/total number person years) x 100,000 = incidence rate per 100,000 person years. In cases where the number of person years in an eligible athlete group is given as a percentage of the total number of person years included in the study, the percentage was used to calculate the number eligible for our study.

Independent dual screening and selection of studies was performed by a team of four reviewers (AL, NP, CM, VL). The primary author (AL) screened all abstracts/titles, and full text articles, and was involved in all resolutions of disagreements at each stage of study selection. 100% of abstracts and titles were dual screened. 90% of full text articles were reviewed by two reviewers due to resources and timing. In cases where multiple reports were published on one data set, only the most recent or complete report was included. In cases where different authors used the same databases, the article with the most complete overall database was included. Authors were contacted electronically with questions about screened texts or with requests for eligible data.

### Data Extraction and Quality Assessment

Data extraction, and evaluation of risk of bias (ROB) was performed in all eligible articles by the lead author (AL); 21 of the 40 eligible articles were independently assessed and extracted by a second person (NP, VL, CM), the remaining 19 individually assessed and extracted articles were checked by a senior author (DN). In cases of disagreement, these were resolved by discussion of the two involved parties. ROB was assessed with a validated tool developed to assess prevalence studies ^29^, and customized for the purposes of this review. We considered studies done prospectively as the highest level of evidence. Retrospective projects done on databases with largely prospectively collected data were also considered higher quality evidence. Both the ROB tool, and the data extraction guide, are included in the supplemental appendix.

### Data Synthesis and Analysis

The primary outcome was the reported incidence rate of SCA and SCD in competitive athletes, and active duty military members age 40 and under. Competitive athletes were considered those participating in athletics of any form or level, including scholastic, recreational, club, collegiate/university level, and professional in or out of season. Active duty military members were included due to the high level of activity that is required of them, the similar age profile, and interest in preventing SCA/D. Pre-specified subgroup analysis was planned by age, sex, race, and by type of sport or military member, as well as level of sport (definitions provided in subgroup analysis in supplementary appendix). Analysis was performed with the Metafor package ^30^ in the R statistical software^31^. A random-effects, generalized linear mixed model was used for meta-analysis. Summary findings are presented as events of SCA and SCD per 100,000 person years with 95% confidence intervals.

Heterogeneity was assessed with *I*^2^ and Chi^2^; pre-specified levels of heterogeneity for *I*^2^ were: <30% considered low; 30-70% considered moderate; and >70% was considered high. A p-value of 0.10 or lower for Chi^2^ statistic was indicated statistical heterogeneity. Assessment of publication bias of the studies included was planned using funnel plots when 10 or more studies were pooled. Sensitivity analyses were planned, if necessary, to evaluate heterogeneity within the results.

### Ethical Approval

Ethical approval was not necessary as only publicly available data was included in this review.

### Role of the Funding Source

No funding was received for this project.

## Results

### Study Selection and Characteristics

After removal of 10,780 duplicates, 20,048 records were identified by the search, and 11 further articles were added by hand search. At the time of screening, we were aware of an important unpublished cohort, which we had elected to include in the final analysis after its publication in 2020^32^. 20,060 abstracts were screened and 323 full texts were assessed for inclusion, of which 40 studies met the criteria (Figure 1). The characteristics of the included studies are provided in the data supplement with selected details given in table one.

**Figure 1:**
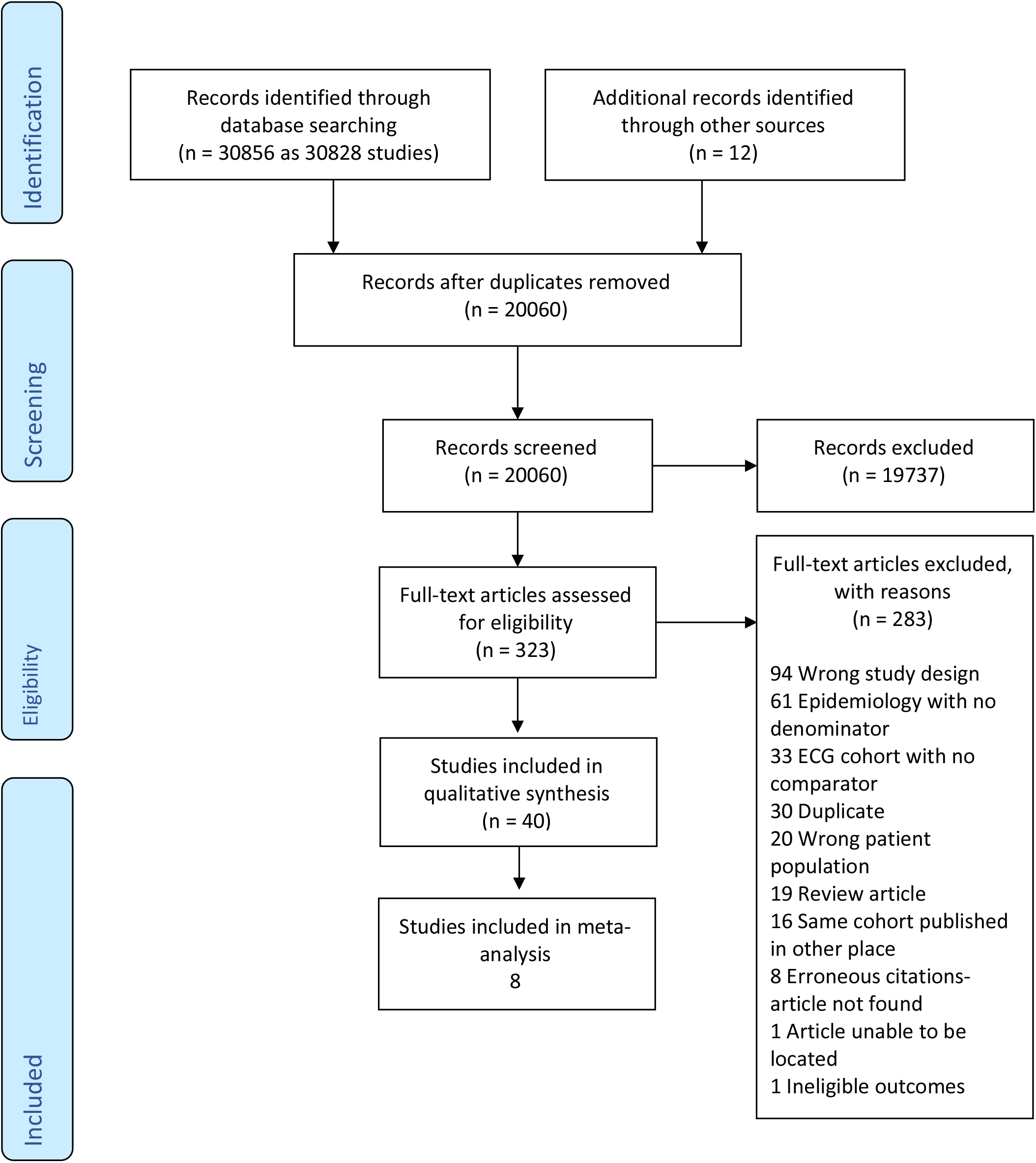
PRISMA flow diagram

34 studies included athletes, ranging from youth to professional levels. Articles included were on athletes from the U.S.(n=16)^6,9,38–42,13,14,32–37^, Europe (n=13)^10,12,51–53,43–50^, International (n=1)^54^ Canada (n=1)^11^, Argentina (n=1)^55^, Israel (n=1)^8^, and Australia (n=1)^56^. Five included papers were performed on military members, four in the United States (U.S.), and one in Finland. ^15–17,57,58^. A study focusing on firefighters in the U.S. ^59^ was also included due to their similarity to military members, and this was grouped with military studies.

25 (61%) included studies employed retrospective cohort designs; 12 were prospective designs (30%) and 3 were surveys (1%), 2 cross-sectional, and one a recurring annual survey. 11 of the above studies reported extractable data only in men ^40,41,43,49,52,56,58,59^. 5 studies were included with a large proportion of the included cohort being men ^15–17,50,57^. Two studies did not have details about gender included in their methodology but were presumed to be either all men, or a high proportion of men ^54,55^. Further details including ages, and levels of sport are included in table one.

### Risk of Bias Assessment and Quality Assessment

12 (30%) studies were determined to present a low overall ROB ^10,11,53,59,12,15–17,32,39,46,50^; 10 (25%) were judged to have moderate risk of bias ^6,9,13,35,37,38,48,49,56,58^; and 18 (45%) were judged to have a high risk of bias ^7,8,44,45,47,51,52,54,55,57,14,33,35,36,40–43^. Of the studies considered low ROB, four pertained to military members. One was judged to be prospective in nature^15^, while the remaining three were retrospective reviews of data collected in a prospective fashion^16,17,59^.

Similarly, the eight articles on athletes include four studies collected in a prospective nature ^1,32,46,50^, with six others considered to be retrospective projects on prospectively collected data^10–12,53^. Further details on ROB judgement on individual studies may be found in table two, with explanations for each study in the supplementary appendix.

### Study Findings and Data Synthesis

The studies included presented substantial clinical, statistical, and methodologic heterogeneity. Incidence levels of SCD ranged from 0.02 to 89.05 per 100,000 person years in included studies (Figure 2). Studies reporting SCA had incidence levels ranged from 0.94 to 63.03 per 100,000 person years (Figure 3).

**Figure 2:**
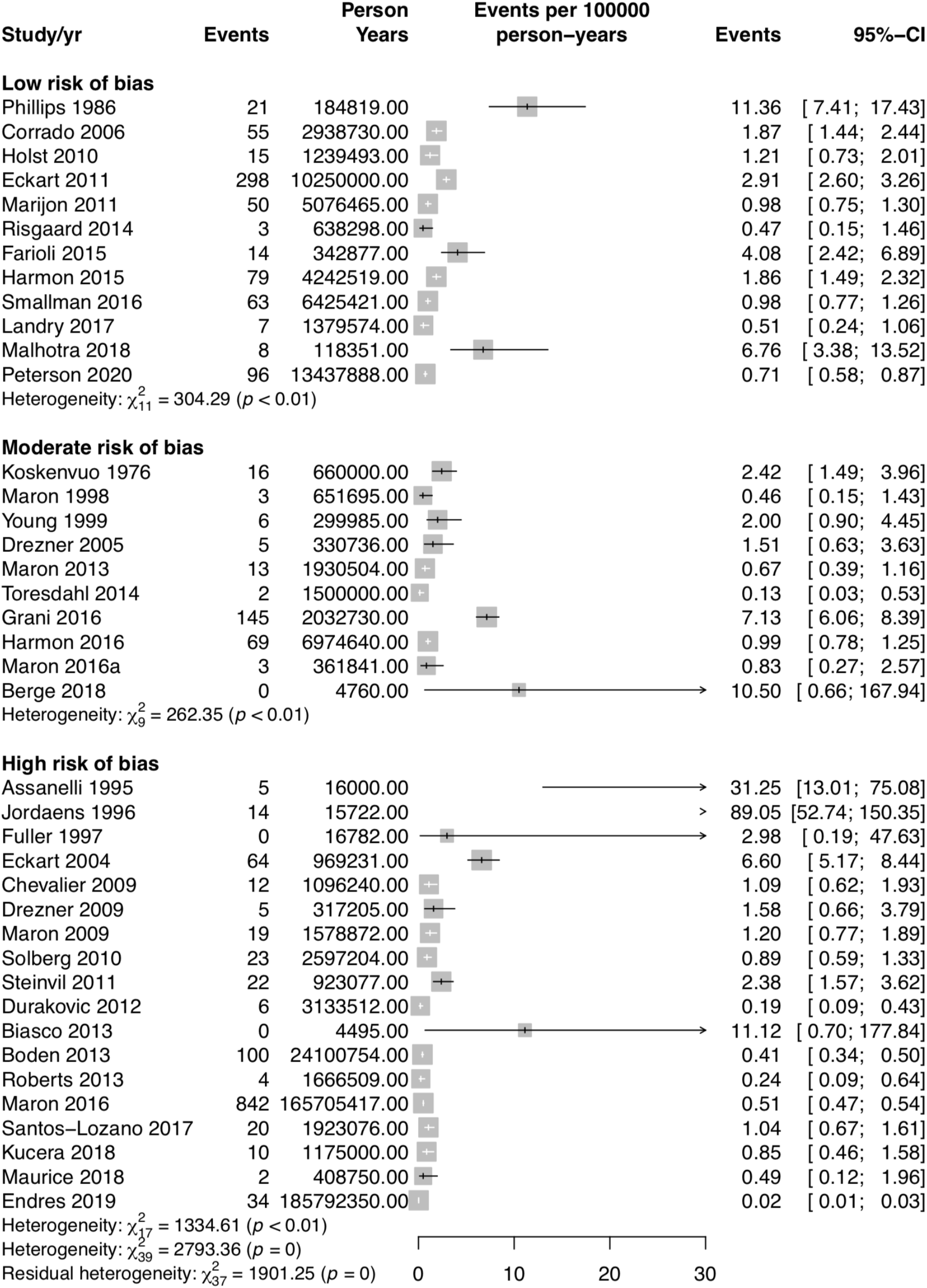
Incidence rates of sudden cardiac death (SCD) reported by included studies.

**Figure 3:**
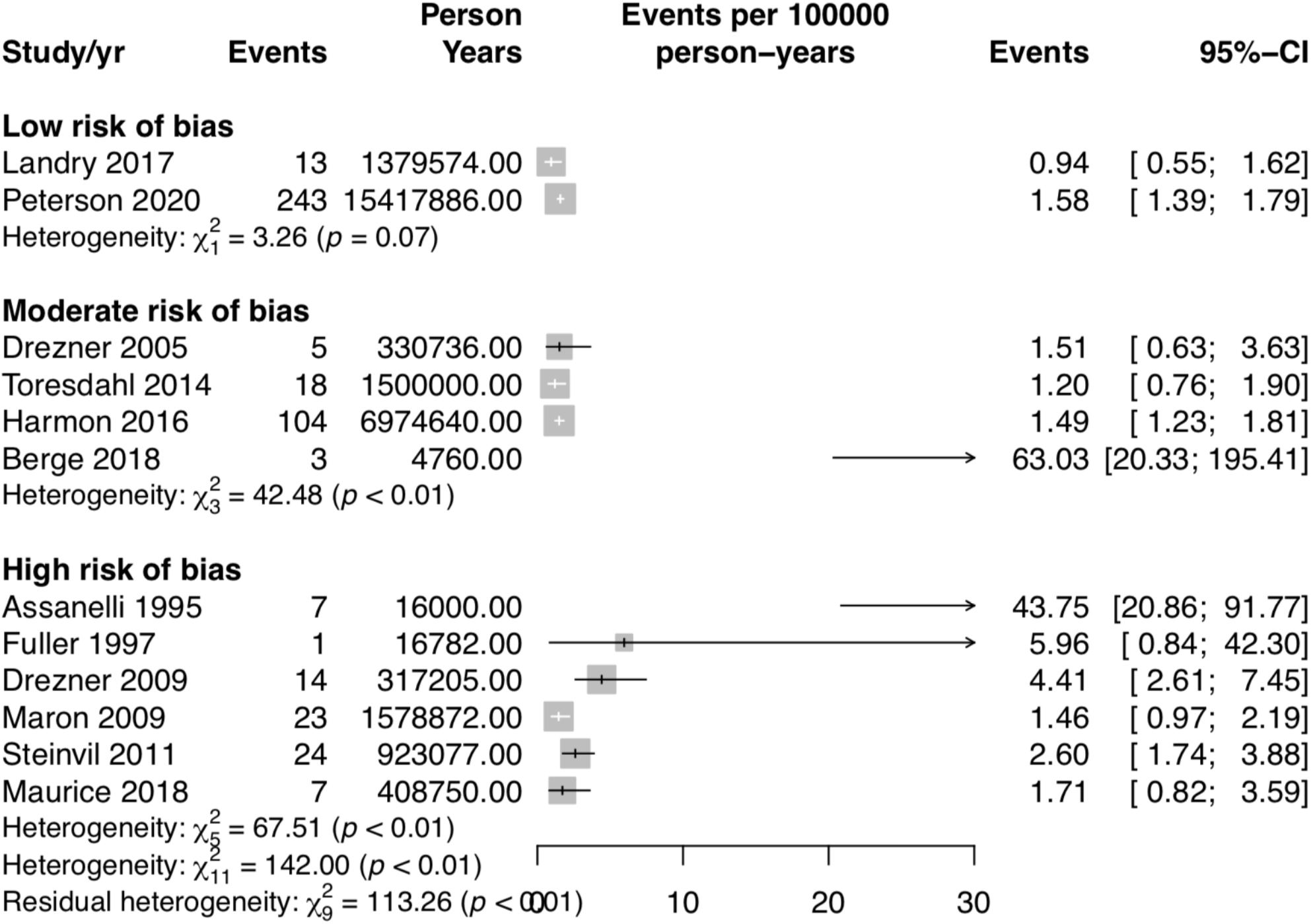
Incidence rates of sudden cardiac arrest (SCA) in reported by included studies

Meta-analysis was performed only in studies considered to be low ROB. Studies including athletes which were collected at a population level^10–12,46,60^ were combined separately from studies focused on broad groups of competitive athletes under 25 years of age^32,39,50^. The population studies had a point estimate of 0.98 [95%CI: 0.62-1.53] SCD per 100,000 athlete years with high heterogeneity (Figure 4). Sensitivity analysis removing Corrado, et. al. (2006) which was an outlier, decreased the heterogeneity in the estimate, from *I*^*2*^ of 78% to 0% and changed the point estimate to 0.90 [95%CI: 0.72, 1.13] SCD per 100,000. The studies which included only competitive athletes were found to have a point estimate of 1.91[95%CI: 0.71; 5.14] SCD per 100,000 with high heterogeneity (Figure 4). It is notable in this meta-analysis that Peterson, et. al. (2020) only provided extractable detail from high school students on SCD events, while providing detail on university and high school athletes for SCA. Sensitivity analysis removing Malhotra et. al (2018) which included primarily male professional soccer players in England, leaving two studies which included a broad sample of both male and female scholastic and university level athletes did not alter the heterogeneity present, but dropped the point estimate to 1.14 [95%CI: 0.59, 2.22].

**Figure 4:**
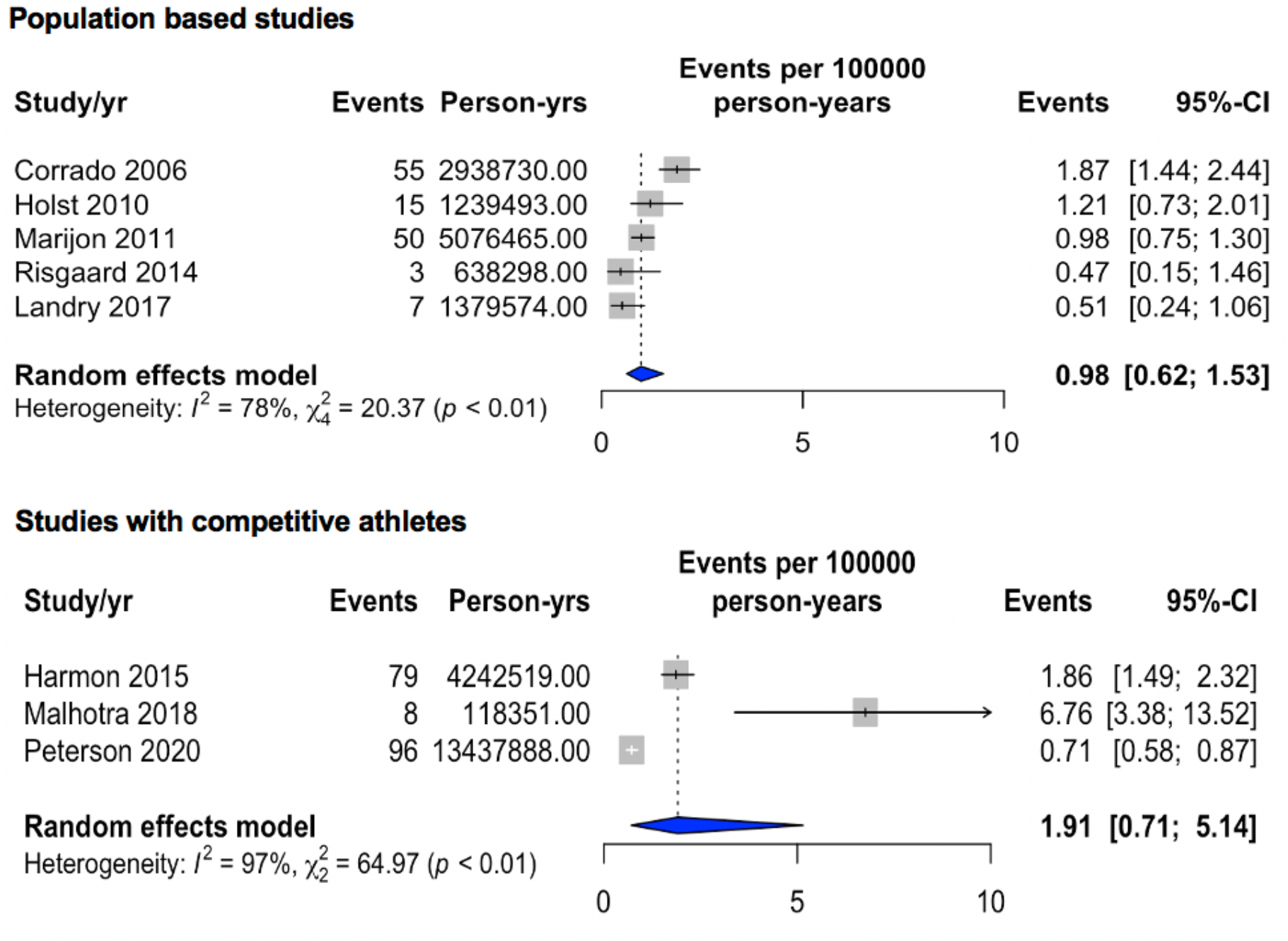
Meta-analyses of incidence rate of sudden cardiac death in low risk of bias studies taking place at population levels, and in studies in competitive athletes between the ages of 14-25.

We elected not to perform synthesis of the low ROB military studies. The populations of two studies^15,16^ included substantial overlap of included subjects, with very different point estimates for SCD 2.91^15^ vs 0.98^16^ per 100,000 military years; the remaining two studies included substantial clinical heterogeneity in the population, with one^17^ dating back nearly 40 years previous to other included studies, and the other including American firefighters^59^. The point estimates of the low ROB studies in the military range from 0.98 in the most recent publication^16^ to 11.36 SCD per 100,000 person years in the oldest^17^.

Inspection of studies reporting SCD in the moderate and high ROB categories largely are in line with the meta-analyses in low ROB studies. 18 of 28 studies report rates under 2.00 per 100,000 athlete years. The remaining ten studies have estimates which range up to 89.05 per 100,000 athlete years. Three of these are studies with zero events, in which estimates should be taken with caution.

There were only two low ROB studies reporting episodes of SCA, and this data was not synthesized. Landry, et. al. is a retrospective population level study in a region of Ontario, Canada, and reported a rate of 0.94 [95%CI: 0.55; 1.62] SCA per 100,000 athlete years; while Peterson, et. al. (2020) was a prospective study reporting SCA in high school and collegiate athletes in the United States reporting 1.58 [95%CI: 1.39; 1.79] SCA per 100,000 athlete years. The moderate and high ROB studies ranged from 1.20 to 63.03 SCA per 100,000 athlete years, with five providing point estimates at less than 2.00, and the remaining five with estimates between 2.00 and 63.03 per 100,000 person years.

Synthesis of subgroups was not performed due to the low number of low ROB studies providing subgroup data. Notable findings from low ROB subgroups indicate higher rates of SCA/D in males compared to females^32,39^ with rates over three times as high in males in both cases.

Separating athletes by race is difficult based on a lack of uniformity in reporting, but generally indicates elevated rates in black athletes in studies comparing high school and collegiate athletes^32,39^ with white, and in one study Hispanic athletes^39^. These findings are similar to studies in the high ROB category reporting on race^34,57^ which similarly show elevated rates in black military members when compared with white counterparts^57^; and white athletes when compared with non-white athletes^34^. Further narrative detail, as well as forest plot figures with results from individual studies on subgroups including by individual sport, and by level of sport is included in the online supplement.

Visual assessment of the funnel plot (included in supplementary appendix) for publication bias showed asymmetry in the direction of larger studies with larger incidence. This finding is likely related to the heterogeneity present in the included studies.

## Discussion

We believe ours to be the first full systematic review on the subject of sudden cardiac arrest and death in athletes, and in military members. Our analysis has shown a substantial amount of heterogeneity within the published literature on the subject, with most papers showing a moderate or high risk of bias. In studies judged to have low ROB, meta-analysis of three published studies in young competitive athletes showed an incidence of SCD to be 1.91[95%CI: 0.71; 5.14] SCD per 100,000; while five studies including large national or regional populations of athletes was found have an incidence of 0.98 [95%CI: 0.62-1.53] SCD per 100,000 athlete years. Few low ROB studies reported extractable SCA details. Landry, et. al. (2017) reported on athletes of all levels in a regional population based study in Ontario, Canada. The point estimate was reported as 0.94 [95%CI: 0.55; 1.62] per 100,000 athlete years. Peterson, et. al. (2020) reported on high school and university athletes with a point estimate of 1.58 [95%CI: 1.39; 1.79] SCA per 100,000 athlete years. Military studies included were not meta-analyzed, but showed a range of estimates of SCD incidence, from 0.98 to 11.36 per 100,000 years.

We believe these findings in high quality studies of athletes confirm the rarity of sudden cardiac arrest and death in athletes. It does appear that the incidence rates in military members may be more frequent than in athletes, but with the heterogeneity present in the estimates, it is difficult to be certain about the point estimate. In both cases, athletes and military, the most extreme estimates come from studies which include populations dating back to the 1970s in the case of Corrado, et. al. (2006), and into the 1960s in the case of Phillips, et. al. (1986). Including these studies when considering the likely true rates of SCD likely unduly influences results when comparing them to more contemporary reports.

Recent high quality estimates in the United States focused on athletes in high school and university include studies which were conducted in a prospective fashion, or reviewed databases which collected information in a prospective manner. In these cases, the estimates of SCD and SCA were under 2 per 100,000^32,39^. In both of these cases, both exertional and non-exertional cases of SCA/D were included in estimates. Criticisms of estimates excluding non-exertional cases have been made in the past^5^, based on the belief that counting on exertional deaths underestimates the risks of SCA/D in the active young population. The same concerns about underestimating risk have been applied to the concept of counting only deaths, while not including arrests in which the subject survives. The estimates provided by these two studies include broad populations of male and female athletes, as well as multi-ethnic studies. They do confirm that the rates of SCA/D in the population of competitive scholastic and university level athletes are greater than other estimates of less than 1 per 100,000^3^, therefore, there is not a reason to believe that the rates in these cases are significantly undercounted.

When considering the results of low ROB studies focused on athletes participating in regular practice and competition, the estimate of SCD is slightly higher than in the population based studies including athletes of all levels, including recreational. Of the population based studies, four took place in European countries, and one in Ontario, Canada. The explanations for this difference are likely multifactorial. Of the population based studies, only Corrado, et. al. (2006) includes all deaths in athletes, rather than exertional deaths only. Corrado^46^ however, focuses more on competitive athletes than the remaining four studies^10–12,53^, and as mentioned previously, dates back to the 1970s for its data collection. It is also true that the population of black athletes is likely to be lower in Europe and Canada, and this must be considered when making comparisons to the United States if the rates in black athletes, as appears to be the case, are considerably higher. It is also notable that the ages included in the population level studies extend into the 30s, which one would expect might increase the frequency of SCD related to coronary artery disease, when compared to other conditions causing SCD in the younger athlete^32^. This fact does not appear influential enough to raise the rates of those seen in competitive athletes.

While the overall rate of SCA/D appear relatively low, attention to subgroups of athletes suggests that there are likely areas where rates are consistently much higher. The rates reported by the authors particularly in black males in the sports of basketball and American football are considerably different than their population estimates. Harmon, et. al. (2015) report a rate of 1 SCD in black male basketball players per 4,380 athlete years at the elite university level, while Peterson, et. al. (2020) report a rate of sudden cardiac arrest and death of 1 per 4,810 athlete years in university level athletes at all levels, and 2,087 in athletes at the elite division one level; and reported a rate of 1 per 28,061 in black American football athletes at all levels, and 1 per 18,031 at the division one level. There are four articles reporting outcomes specifically in soccer athletes^49,50,52,54^, and two reporting subgroups of soccer athletes^32,39^. The high quality articles reporting SCD events report rates of 4.22 in university athletes in the United States, and 6.76 in professionals in England per 100,000 athlete years, while the reports of SCA in university athletes in the United States are far lower in the Peterson, et. al. 2020 article, at 1.28 SCA per 100,000 athlete years. The variability in the results of these studies makes it somewhat difficult to interpret, or understand the true risk associated with soccer, in comparison to other sports.

Applying the data presented in this systematic review confirms the general understanding that occurrence in the young active population for sudden cardiac death appears to be low. This information is further able to inform decisions and recommendations on screening the young active population prior to their participation in sport, or vigorous physical activity such as military service. As is noted in the recent recommendations in the United Kingdom^20^, the rarity in the population of the events make screening and primary prevention a difficult task. It is notable that in the two studies reporting on data collected on cohorts of soccer athletes that had undergone screening ECG, both report rates higher than seen in overall populations, Malhotra, et. al. (2018) reports eight events of SCD for a rate of 6.76 [95%CI: 3.38, 13.52] in a low ROB study; while Berge, et. al. (2018) reports 3 events of SCA for a rate of 63.03 [95%CI: 201.33; 195.41] in a study judged to be of moderate ROB related to its use of media sources, and low number of overall athlete years. It is important to note that two of the deaths reported in Malhotra, et. al. were in athletes identified at high risk for SCA/D, who continued to participate against medical advice. While imprecise, and probably not large enough cohorts to settle the question certainly, these two articles give pause to the idea that ECG screening lowers incidence of SCA/D. In addition to data presented over the past decade about the poor sensitivity to identify conditions which put one at risk of SCA/D with history and physical alone^61,62^, the utility of screening to prevent these events remains unproven.

We believe the primary strength of our review lies in the breadth of the search, leading to the ability of this review to draw data from multiple, large population level studies, as well as studies focusing specifically on scholastic and college athletes, and studies focusing on military members. We believe this review provides the most up-to-date and precise estimates of population level incidence including athletes of all types, as well as incidence in competitive athletes. We also believe narrative review of subgroups adds to the overall knowledge base about those with high and low risk levels. This review also provides an assessment of the level of evidence published on the subject of the incidence of SCA/D.

Limitations of our review include substantial clinical and statistical heterogeneity in the published data, as well as the overall low proportion of data judged to have low ROB. Many of the studies included only males, and/or single sports, and were judged to be at moderate or high risk of bias, tempering the robustness of their findings. Most of the data included in this review is drawn from studies in Europe and the United States, potentially limiting the applicability of our findings outside of these regions. Due to available resources and timing, not all included studies were double extracted or assessed for risk of bias.

## Conclusion

This systematic review provides the most comprehensive review of the available data on sudden cardiac arrest and death in active populations under the age of 40. Our estimate of the incidence of SCD in high quality studies shows SCD to be a rare occurrence in both population level studies including all levels of athletes, 0.98 [95%CI: 0.62-1.53] SCD per 100,000 athlete years, and focused studies evaluating young competitive athletes between the ages of 14 and 25, 1.91[95%CI: 0.71; 5.14] SCD per 100,000. Our evaluation of the published literature suggests higher incidence of both SCD and SCA with male sex, black athletes, and potentially in the military compared with athletic involvement. We believe that the estimates presented here for broad populations of competitive athletes are not likely to be changed by further publications. We do believe that population based studies may be improved upon by including events at all times, in comparison to events which occur only with exertion. As has been suggested previously^63^, a uniform reporting system of sudden deaths in the young active population would benefit the understanding of this condition, and advance a more reliable understanding of the incidence of both sudden cardiac arrest and death.

## Data Availability

Reasonable requests for data will be considered by email request to first author.

---

**Table 1:** Characteristics of included studies

| 1st Author and year of publication | Methodology | Open vs closed cohort | Exertional only or all cardiac deaths | Method of case identification | SCA or SCD Reported | Category of sport or activity | Level of sport or activity | ECG screened | Ages (yrs) | Sex | Study Length (yrs) | ROB |
| --- | --- | --- | --- | --- | --- | --- | --- | --- | --- | --- | --- | --- |
| Phillips 1986 | Retrospective Cohort | Known | Exertional | Autopsy reports | SCD only | Military | Military | No | 17-28 | M & F | 6 | Low |
| Corrado 2006 | Prospective Cohort | Open | All | Combined media reports/autopsy and medical records | SCD only | Multiple | Competitive | Portion | 12-35 | M & F | 24 | Low |
| Holst 2010 | Retrospective Cohort | Open | Exertional | Death certificate | SCD only | Soccer | Competitive | Not described | 12-35 | M & F | 12 | Low |
| Eckart 2011 | Prospective Cohort | Known | All | Combined autopsy/other official record such as medical, military, athletic body | SCD only | Military | Military | No | 18-34 | M & F | 1 | Low |
| Marijon 2011 | Retrospective Cohort | Open | Exertional | Combined media reports/autopsy and medical records | SCD (SCA included in SCD count) | Multiple | Competitive | No | 10 -35 | M & F | 26 | Low |
| Risgaard 2014 | Retrospective Cohort | Open | Exertional | Combined media reports/autopsy and medical records | SCD only | Multiple | Competitive | No | 12-35 | M & F | 9 | Low |
| Farioli 2015 | Retrospective Cohort | Open | All | Combined autopsy/other official record such as medical, military, athletic body | SCD only | Military | Military | No | 18-34 | M | 25 | Low |
| Harmon 2015 | Prospective Cohort | Open | All | Combined media reports/autopsy and medical records | SCD only | Multiple | University | Not described | 17-24 | M & F | 26 | Low |
| Smallman 2016 | Retrospective Cohort | Known | Exertional | Combined autopsy/other official record such as medical, military, athletic body | SCD only | Military | Military | No | <35 | M & F | 10 | Low |
| Landry 2017 | Retrospective Cohort | Open | Exertional | Combined autopsy/other official record such as medical, military, athletic body | SCA and SCD | Multiple | Multiple | Not described | 12-35 | M & F | 32 | Low |
| Malhotra 2018 | Prospective Cohort | Known | All | Combined autopsy/other official record such as medical, military, athletic body | SCD only | Soccer | Elite | Yes | 15-17 at time of screen | M & F | 15 | Low |
| Peterson 2020 | Prospective Cohort | Open | All | Combined autopsy/other official record such as medical, military, athletic body | SCA and SCD | Multiple | Multiple | Not described | 11-29 | M & F | 4 | Low |
| Koskenvuo 1976 | Retrospective Cohort | Known | Not described | Combined autopsy/other official record such as medical, military, athletic body | SCD only | Military | Military | No | 18-24 | M | 6 | Moderate |
| Maron 1998 | Retrospective Cohort | Open | Exertional | Autopsy reports | SCD only | Multiple | Scholastic | No | 13-19 | M & F | 1 | Moderate |
| Young 1999 | Retrospective Cohort | Open | Exertional | Autopsy reports | SCD only | Australian football | Competitive | No | 15-37 | M | 3 | Moderate |
| Drezner 2005 | Survey | Open | Not described | Facility event report (ex: school records) | SCA and SCD | Multiple | Elite | Not described | 17-24 | M & F | 0.5 | Moderate |
| Maron 2013 | Retrospective Cohort | Open | All | Combined media reports/autopsy and medical records | SCD only | Multiple | Scholastic | No | 12 -18 | M & F | 3 | Moderate |
| Toresdahl 2014 | Prospective Cohort | Known | All | Interview family/eyewitness | SCA and SCD | Multiple | Scholastic | No | 14-19 | M & F | 11 | Moderate |
| Harmon 2016 | Prospective Cohort | Open | All | Media reports | SCA and SCD | Multiple | Scholastic | No | 14-19 | M & F | 10 | Moderate |
| Grani 2016 | Retrospective Cohort | Open | Exertional | Autopsy reports | SCD only | Multiple | Multiple | Portion | 10-39 | M | 7 | Moderate |
| Maron 2016a | Retrospective Cohort | Open | All | Autopsy reports | SCD only | Multiple | Multiple | No | 14-23 | M & F | 20 | Moderate |
| Berge 2018 | Retrospective Cohort | Known | All | Media reports | SCA and SCD | Soccer | Elite | Yes | 18-40 | M | 15 | Moderate |
| Assanelli 1995 | Prospective Cohort | Not Described | Not described | Medical records | SCA and SCD | Multiple | Multiple | Yes | unclear | M & F | 5 | High |
| Jordaens 1996 | Retrospective Cohort | Open | Not described | Media reports | SCD only | Cycling | Elite | Not described | unknown | M & F | 2 | High |
| Fuller 1997 | Prospective Cohort | Known | Not described | Not reported | SCA and SCD | Multiple | Scholastic | Yes | <20 | M & F | 8 | High |
| Eckart 2004 | Retrospective Cohort | Known | All | Combined autopsy/other official record such as medical, military, athletic body | SCD only | Military | Military | No | 17-35 | M & F | 3.3 | High |
| Chevalier 2009 | Prospective Cohort | Open | Exertional | Medical records | SCD only | Multiple | Multiple | No | < 35 | M & F | 21 | High |
| Drezner 2009 | Survey | Open | Not described | Facility event report (ex: school records) | SCA and SCD | Multiple | Scholastic | No | 14-19 | M & F | 10 | High |
| Maron 2009 | Retrospective Cohort | Open | All | Combined media reports/autopsy and medical records | SCA and SCD | Lacrosse | Competitive | No | 14-23 | M & F | 32 | High |
| Solberg 2010 | Retrospective Cohort | Open | Exertional | Combined autopsy/other official record such as medical, military, athletic body | SCD only | Multiple | Multiple | No | 15-34 | M | 10 | High |
| Steinvil 2011 | Retrospective Cohort | Open | Not described | Media reports | SCA and SCD | Multiple | Competitive | Portion | unclear | M & F | 15 | High |
| Durakovic 2012 | Retrospective Cohort | Open | Not described | Combined media reports/autopsy and medical records | SCD only | Multiple | Competitive | Portion | 17-29 | M | 8 | High |
| Biasco 2013 | Retrospective Cohort | Known | Not described | Not reported | SCA and SCD | Soccer | Elite | Yes | Mean 23.6 | M | 5 | High |
| Boden 2013 | Prospective Cohort | Open | Not described | Combined eyewitness interview and medical/autopsy record | SCD only | American Football | Multiple | Not described | 14-25 | M | 6 | High |
| Roberts 2013 | Retrospective Cohort | Open | Exertional | Insurance records | SCD only | Multiple | Scholastic | No | 12 -19 | M & F | 19 | High |
| Maron 2016 | Retrospective Cohort | Open | All | Combined media reports/autopsy and medical records | SCD only | Multiple | Competitive | Not described | 15-24 | M & F | 10 | High |
| Santos-Lozano 2017 | Retrospective Cohort | Open | Exertional | Media reports | SCD only | Soccer | Elite | Portion | 16-34 | Not reported | 21 | High |
| Kucera 2018 | Prospective Cohort | Open | All | Combined eyewitness interview and medical/autopsy record | SCD only | American Football | Competitive | No | <40 | M | 25 | High |
| Maurice 2018 | Survey | Open | Exertional | Facility event report (ex: school records) | SCA and SCD | Rugby | Competitive | No | unknown | M & F | 1 | High |
| Endres 2019 | Retrospective Cohort | Open | Exertional | Media reports | SCD only | Multiple | Multiple | No | 6-17 | M & F | 27 | High |

SCD=Sudden cardiac death, SCA=Sudden cardiac arrest, M=Male, F=Female
Athlete categories: Elite=professional or Division 1 NCAA athletes in U.S.; Competitive= athletes participating in organized sport with practices and competitions; University=athletes competing in all levels at a university setting; Scholastic=competing for schools between middle and high school equivalents in the U.S. (approximately ages 12-19); Multiple=combination of different levels;
Military=active duty military, and one study including firefighter

**Table 2:**
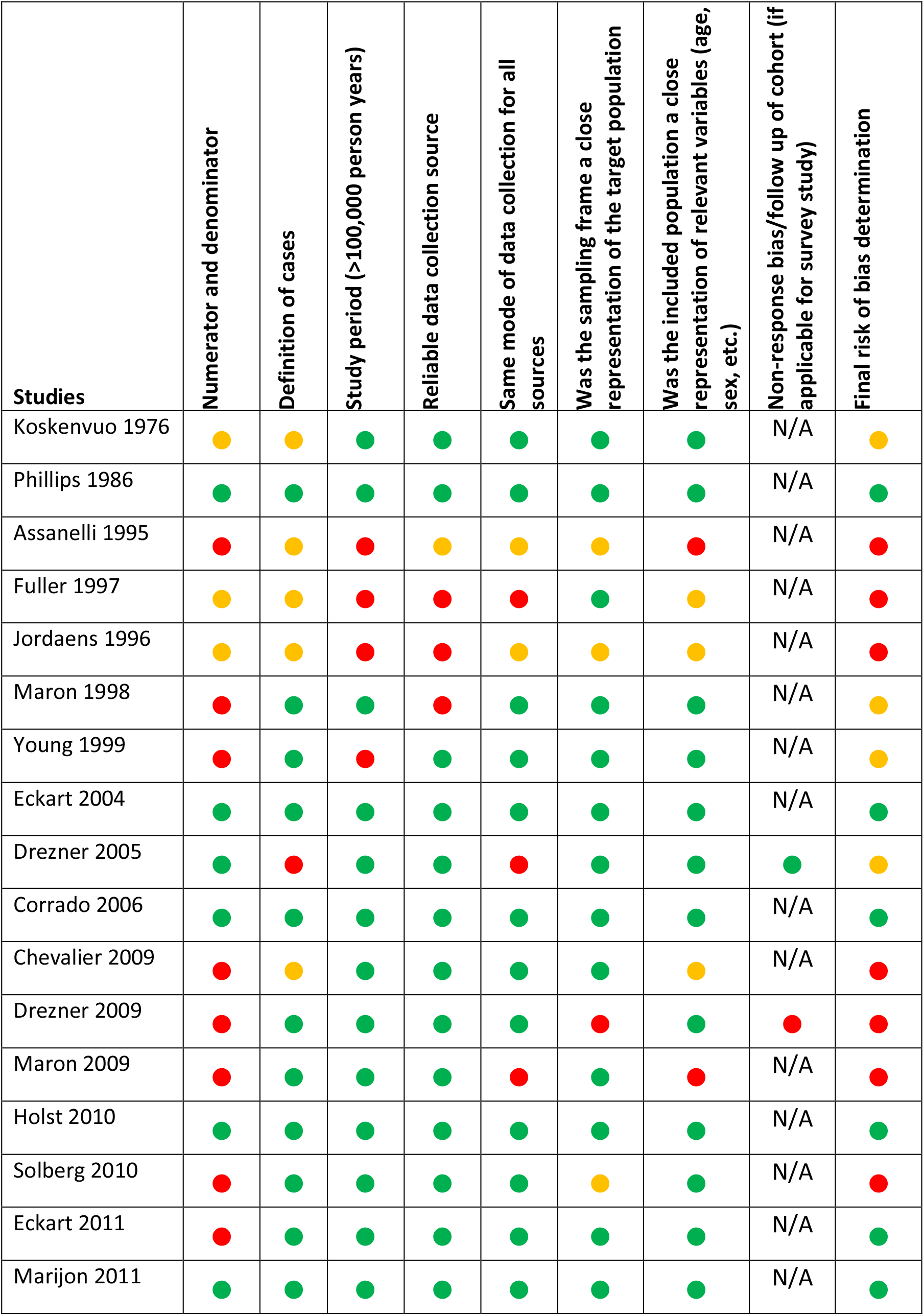
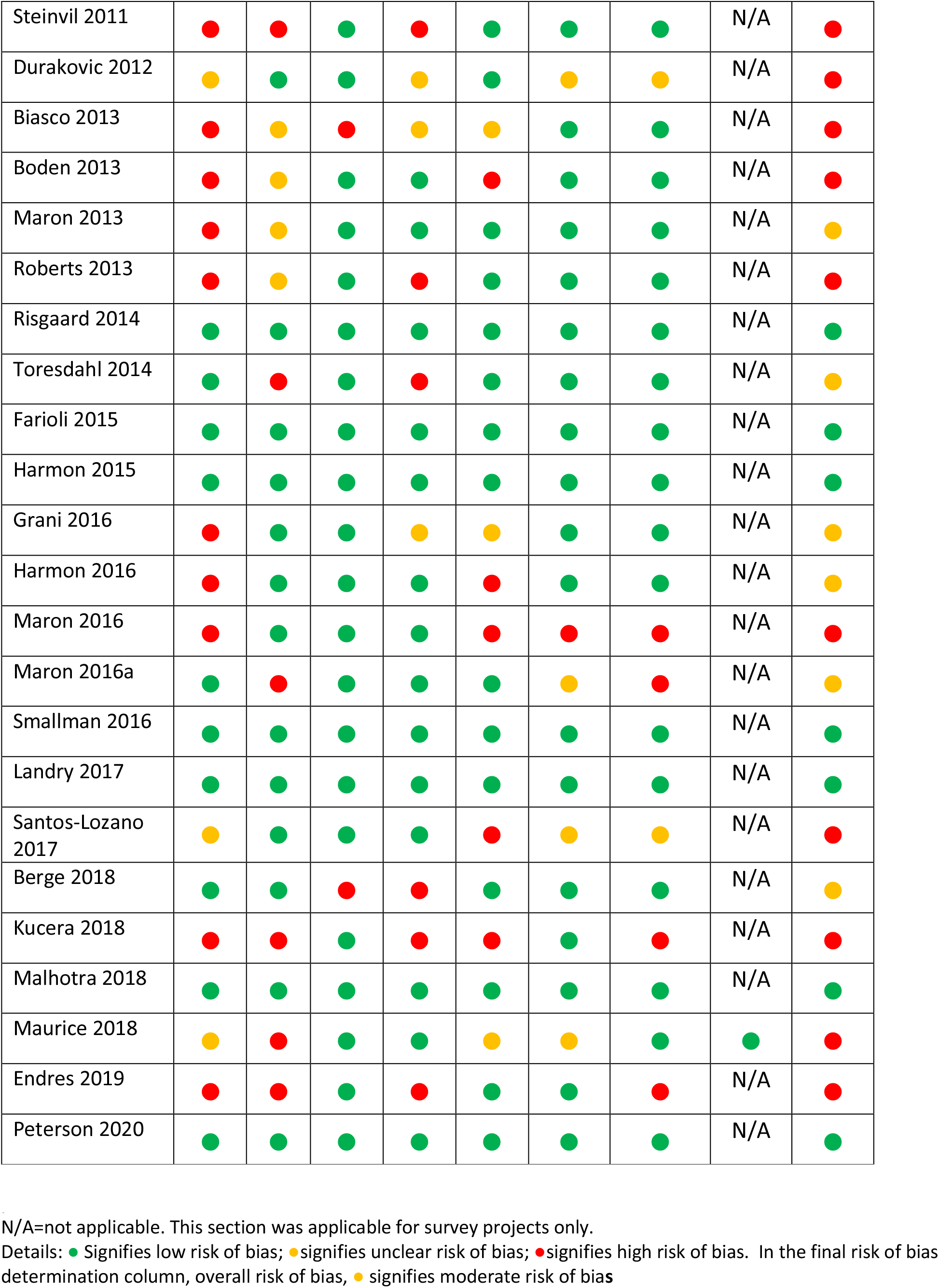
Risk of bias table for included studies reporting on incidence based on customized version of risk of bias in prevalence studies tool ^29^.

## Notes

### Competing Interest Statement

The authors have declared no competing interest.

### Clinical Trial

PROSPERO: CRD42019125560

### Author Declarations

Systematic review. IRB exempt.

